# Joint association of C-reactive protein-triglyceride glucose index-frailty index and non-exercise estimated cardiorespiratory fitness with all-cause mortality in adults aged ≥ 45 years with cardiovascular-kidney-metabolic syndrome stages 0–3: a cross-cohort study using NHANES and CHARLS

**DOI:** 10.64898/2026.06.16.26355835

**Authors:** Jie An, Qianhua Feng, Jie Li, Ya Luo, Meng Yu, Min Xu, Dandan Yang, Qiang She

## Abstract

**Background:** The joint association of the C-reactive protein-triglyceride glucose index-frailty index (CTI-FI) and non-exercise estimated cardiorespiratory fitness (eCRF) with all-cause mortality (ACM) in adults aged ≥45 years with cardiovascular-kidney-metabolic (CKM) syndrome stages 0–3 remains unexplored.

**Methods:** Participants were enrolled from the National Health and Nutrition Examination Survey (NHANES; derivation cohort) and the China Health and Retirement Longitudinal Study (CHARLS; external validation). Covariate selection was performed using LASSO regression. Weighted Cox models were applied across four adjustment models to evaluate the independent associations of CTI-FI and eCRF with ACM. Dose-response patterns were examined with restricted cubic splines (RCS). Subgroup, sex-stratified, and mediation analyses tested robustness and pathways.

**Results:** A total of 6,662 participants from NHANES (median follow-up 10 years; 1,276 ACM, 19.2%) and 3,418 participants from CHARLS (9 years; 391 deaths, 11.4%) were included. Per 1-unit increase in CTI-FI, the risks increased by 44% for ACM (HR 1.44; 95% CI 1.31–1.57) and by 54% for cardiovascular mortality (CVM, HR 1.54; 95% CI 1.33–1.79); per 1-MET increase in eCRF, the risks decreased by 10% (HR 0.90; 95% CI 0.85–0.94) and by 18% (HR 0.82; 95% CI 0.75–0.90), respectively (all P < 0.001). Compared with the low CTI-FI + high eCRF group, the high CTI-FI + low eCRF group was associated with a significantly higher risk of ACM (HR 2.74; 95% CI 2.20–3.40) and CVM (HR 5.04; 95% CI 3.02–8.40). RCS analysis showed a nonlinear CTI-FI- ACM association. The model with CTI-FI and eCRF achieved a C-index of 0.78 for ACM and 0.83 for CVM. CTI-FI and eCRF bidirectionally mediated each other’s associations with ACM and CVM. Specifically, eCRF accounted for 16.4%–23.5% of CTI-FI-related mortality risk, whereas CTI-FI accounted for 23.9%–32.2% of eCRF’s survival benefit (all P < 0.001).

**Conclusions:** Higher CTI-FI and lower eCRF independently and jointly predict increased mortality, with bidirectional mediation indicating that improving one may partially offset the adverse effect of the other. These findings highlight potential therapeutic targets for early CKM syndrome management.

**Graphical abstract:** 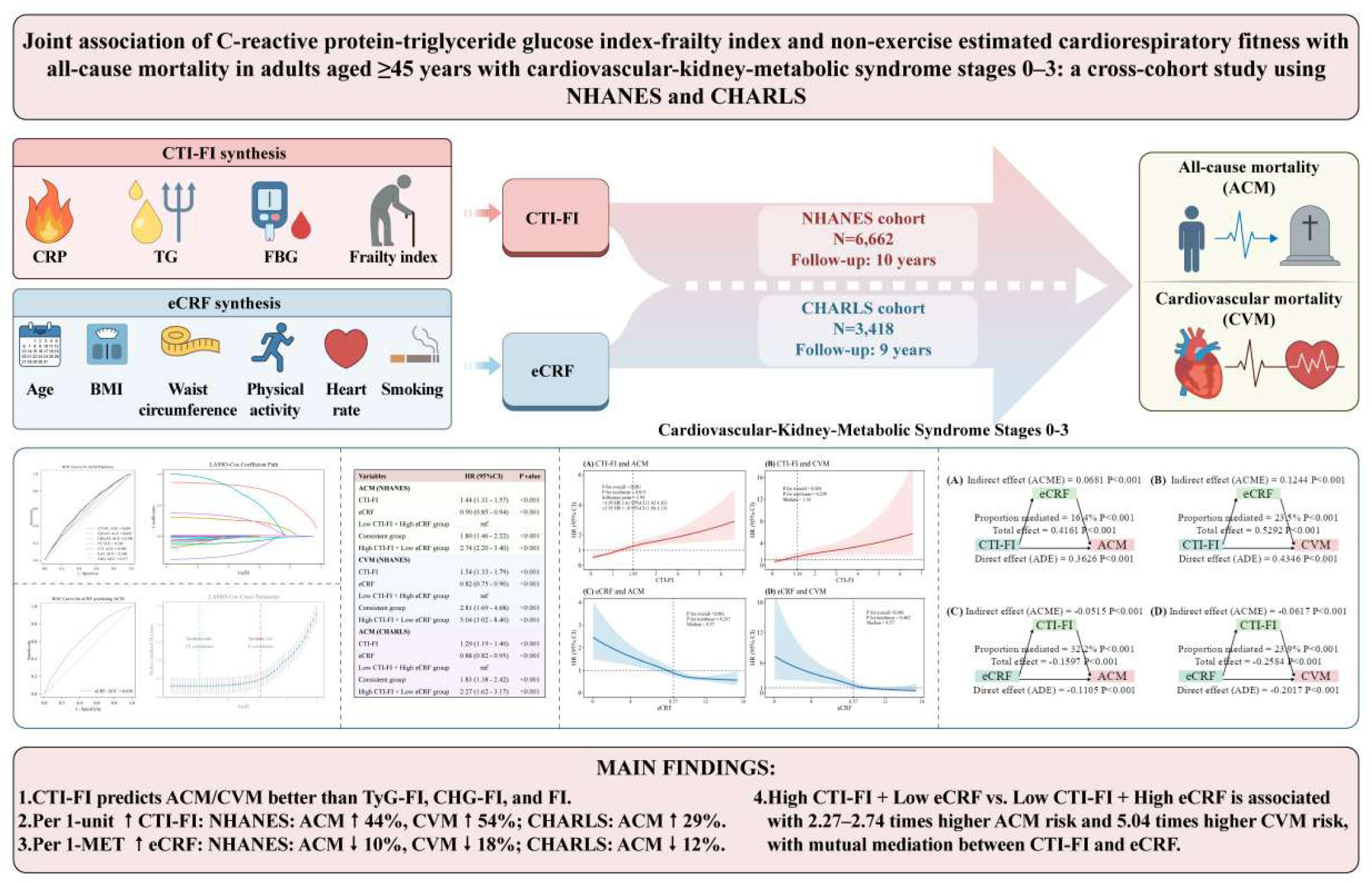

**Research Insights:** *What is currently known about this topic?:* CKM syndrome is common but lacks early risk stratification. CTI-FI and eCRF each predict mortality in general populations, yet their independent and joint associations with mortality in CKM stages 0-3 remain uncharacterised.

*What is the key research question?:* Are CTI-FI and eCRF independently and jointly associated with all-cause and cardiovascular mortality in adults ≥45 years with CKM stages 0-3, and are findings consistent across US and Chinese cohorts?

*What is new?:* Per 1-unit increase was associated with a 44% higher risk of ACM and a 54% higher risk of CVM. Per 1-MET higher eCRF, ACM and CVM risks were reduced by 10% and 18%, respectively. Compared with the Low CTI-FI + High eCRF group, the High CTI-FI + Low eCRF group had a 2.74-fold higher risk of ACM and a 5.04-fold higher risk of CVM. Bidirectional mediation: eCRF explained 16-24% of CTI-FI-related mortality risk, while CTI-FI explained 24-32% of eCRF’ s survival benefit. Dose-response and mediation patterns differed by sex and population.

*How might this study influence clinical practice?:* This study introduces two non-exercise, easily ascertainable indices—CTI-FI and eCRF—for early risk stratification in adults with CKM stages 0-3. The bidirectional mediation highlights that improving either metabolic-inflammatory status (CTI-FI) or cardiorespiratory fitness (eCRF) could partially counteract the adverse effect of the other, informing targeted lifestyle and pharmacological interventions.

## Backgrounds

Cardiovascular-kidney-metabolic syndrome (CKM) represents a highly prevalent systemic disorder in which metabolic abnormalities, chronic kidney damage, and cardiac structural/functional alterations interact and drive each other forward.^1, 2^ Accumulated evidence highlights insulin resistance (IR), chronic low-grade inflammation, oxidative stress, and endothelial dysfunction as core pathophysiological mechanisms underlying CKM pathogenesis and progression.^3,4^ Because CKM affects a large fraction of the population and substantially elevates the risk of all-cause mortality (ACM) as well as adverse cardiovascular events, it places a heavy burden on healthcare systems worldwide.^5–7^ Therefore, the American Heart Association (AHA) has underscored that CKM stages 0-3 constitute a critical early intervention window, in which timely risk stratification and targeted management may attenuate end-organ damage and improve long-term survival. Nevertheless, the absence of accessible early biomarkers for rapid population screening remains a major obstacle to the early identification and intervention of CKM. Accordingly, there is an unmet clinical demand for simple, feasible, and broadly applicable prognostic tools to refine risk evaluation and optimize individualized CKM management.^8^

The triglyceride-glucose index combined with frailty index (TyG-FI) has recently been proposed as a prognostic indicator for stroke, cardiometabolic multimorbidity (CMM) and ACM in patients with CKM stages 0–3.^9–11^ The combined cholesterol, high-density lipoprotein, glucose, and frailty index (CHG-FI) has similarly been shown to exhibit strong predictive value for adverse outcomes in established CMM, even outperforming TyG-FI in some analyses.^12^ Yet, both indices are limited by their omission of chronic inflammation. To address this gap, the C-reactive protein-triglyceride glucose index- frailty index (CTI-FI) has been proposed, integrating inflammation, insulin resistance (IR), and physiological reserve into a single composite biomarker. A recent study has confirmed that CTI-FI is a predictor of new-onset stroke.^13^ However, their prognostic utility in the early to intermediate stages of CKM (stages 0-3) is yet to be elucidated. More importantly, no head-to-head comparison has been made to evaluate the relative or joint predictive performance of CTI-FI, TyG-FI, and CHG-FI for ACM in this specific population. Given that CKM stages 0-3 represent a window for halting disease progression, identifying the most informative composite biomarker is of paramount clinical importance.

Beyond these metabolic frailty indicators, another core cardiovascular metric warrants consideration: cardiorespiratory fitness (CRF), a physiological index that reflects the body’s ability to take up and utilize oxygen, has emerged as a powerful physiological indicator of overall cardiovascular and metabolic health.^14–16^ Consistent epidemiological findings support a protective role of greater CRF against ACM, a pattern observable in the general population alongside individuals with obesity, diabetes mellitus, and diabetic kidney disease (DKD).^17–21^ Although exercise-based cardiopulmonary exercise testing serves as the gold standard for CRF evaluation, it is limited by complex procedures, multiple contraindications, and poor feasibility for large-scale screening, particularly in older adults with multimorbidity.^22^ By contrast, non-exercise estimated cardiorespiratory fitness (eCRF), derived from routine clinical variables and highly concordant with directly measured CRF, overcomes these obstacles. Importantly, longitudinal cohort studies have shown that each 1-MET increment in eCRF corresponds to a 7%–41% reduction in ACM and a 10%–45% reduction in cardiovascular mortality (CVM), highlighting its robust prognostic discrimination and potential for broad clinical implementation.^15,23^

To date, however, no study has evaluated the individual prognostic value of CTI-FI or eCRF for ACM in adults aged ≥45 years with CKM stages 0–3, let alone their joint association. Given that inflammation, insulin resistance, frailty, and low fitness often co-exist and may synergistically amplify risk, combined assessment could identify high-risk individuals missed by single-factor evaluation. Furthermore, the bidirectional mediation between CTI-FI and eCRF has never been explored. For instance, does low eCRF mediate excess risk linked to elevated CTI-FI, or does high CTI-FI attenuate the protective survival effects of eCRF? Answering these questions could shift management toward integrated multi-domain intervention. It also remains unclear whether adding CTI-FI and eCRF improves risk reclassification. Therefore, this cross-cohort study aimed to evaluate independent/joint associations of CTI-FI and eCRF with ACM and CVM; to examine dose-response patterns; to quantify bidirectional mediation; and to assess added predictive value. By addressing these objectives, we aim to provide a low-cost, scalable risk toolkit for early CKM management.

## Method and materials

### Study Design and Population Source

Using a longitudinal cohort design with NHANES for model development and CHARLS for external validation, we evaluated the temporal associations of baseline CTI-FI and eCRF with subsequent ACM risk. NHANES provides repeated cross-sectional health examination data from the non-institutionalized US civilian population; CHARLS constitutes a population-based prospective cohort enrolling community- dwelling Chinese adults aged 45 years and older. Both datasets contain detailed information on demographics, lifestyle factors, biochemical measurements, and mortality status. We excluded participants aged <45 years at first, then sequentially ruled out individuals with missing data on key variables including CRP, fasting blood glucose (FBG), triglyceride (TG), high-density lipoprotein cholesterol (HDL-C), body mass index (BMI), waist circumference (WC), resting heart rate, physical activity (PA) and smoking status; participants with outliers, missing CKM stage 4 information or unavailable mortality outcomes were further excluded. Ultimately, the final analytical cohort contained 6,662 participants from the National Health and Nutrition Examination Survey (NHANES, 1999–2010 and 2015–2018, initial N = 81,385) and 3,418 participants from the China Health and Retirement Longitudinal Study (CHARLS, 2011–2012, initial N = 17,705), who were subsequently divided into three subgroups (Low CTI-FI + High eCRF group, Consistent group, High CTI-FI + Low eCRF group), and the detailed sample selection process is illustrated in **Figure. 1**.

**Figure 1.**
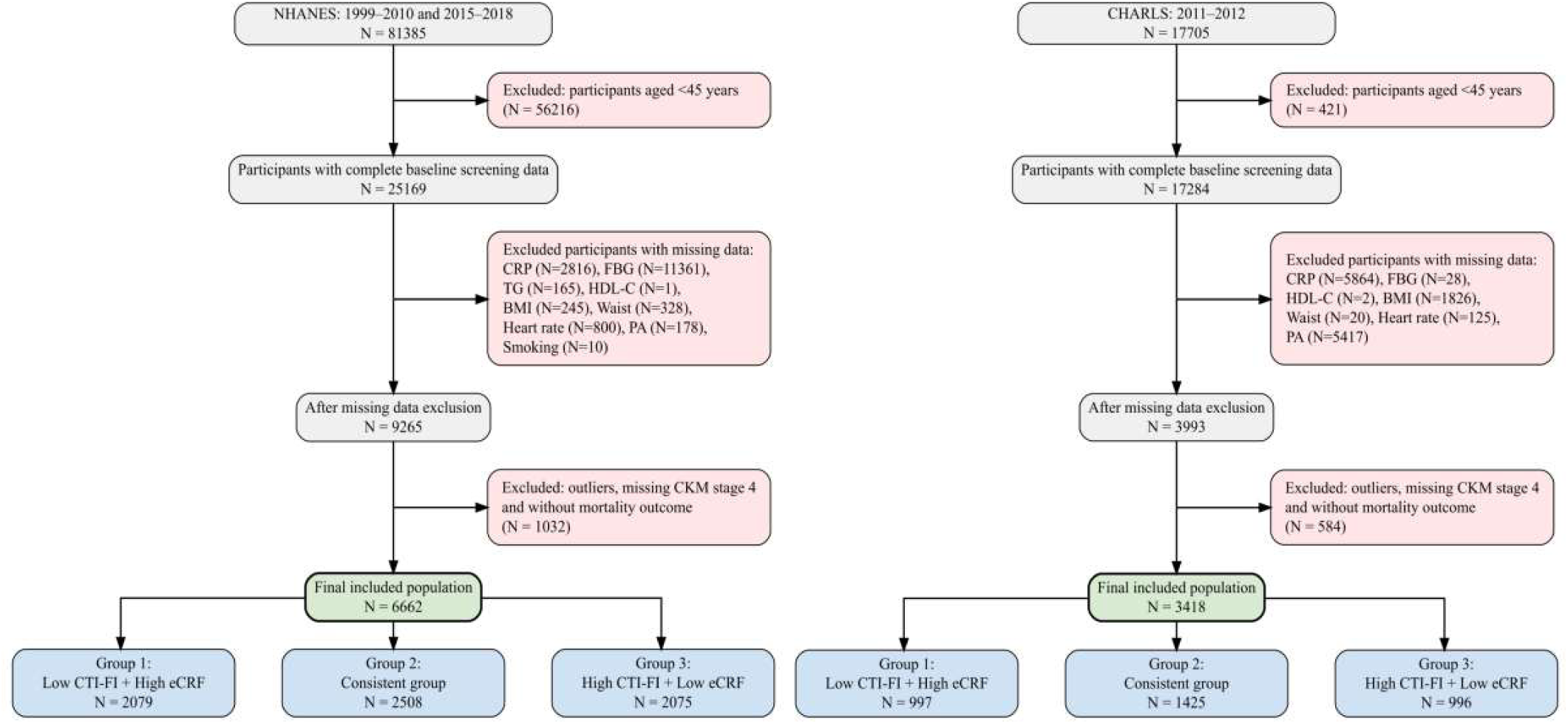
Study flow diagram.

### Data Collection and Variable Definitions

We merged all datasets using unique participant identifiers (SEQN for NHANES, ID for CHARLS). Baseline variables included demographics (age, sex, marital status, education level), lifestyle factors, comorbidities (diabetes, hypertension, chronic kidney disease [CKD]), medication use, CKM stages, standard physiological/laboratory measures, and derived indices (TyG, CHG, CTI, FI, TyG-FI, CHG-FI, CTI-FI, eCRF). Outcomes were ACM and CVM. All biochemical assays followed standardized protocols from the respective surveys. The FI was constructed using the cumulative deficit approach. In NHANES, 49 items were used to calculate the FI; in CHARLS, 32 items were used. Each deficit was dichotomized or polytomized based on clinically meaningful cut-off values, and the FI was computed as the proportion of deficits present. Detailed descriptions of the items and their cut-off values are provided in **Tables S1 and S2**. CKM stages 0-4 were defined as the AHA Presidential Advisory.^1^ Stage 0: No CKM-related risk factors (overweight, abdominal obesity, hypertension, hypertriglyceridemia, prediabetes, diabetes, metabolic syndrome, CKD, or cardiovascular disease [CVD]). Stage 1: Abdominal obesity, overweight, and/or prediabetes, with no other risk factors present. Stage 2: Metabolic disorders (type 2 diabetes, hypertension, elevated triglycerides, metabolic syndrome) or moderate-to-high-risk CKD (G3-G4), without CVD. Stage 3: Presence of subclinical CVD, very-high-risk CKD (stage G4 or G5), or high 10-year predicted CVD risk as assessed by established risk scores. The high atherosclerotic cardiovascular disease (ASCVD) risk threshold was defined as a Framingham Risk Score (FRS) of ≥20% for NHANES participants and as a predicted 10-year ASCVD risk of ≥ 10% for CHARLS participants (**Tables S3, S4A and 4B**).^24,25^ The components of each risk score are detailed in the corresponding supplementary tables. Stage 4: Clinical CVD (coronary heart disease, heart failure, stroke, peripheral artery disease, atrial fibrillation) in combination with metabolic risks or CKD. The estimated glomerular filtration rate (eGFR) was calculated using the CKD-EPI (CKD Epidemiology Collaboration) equation [26]. The calculation formulas for multiple variables are as follows:^10,12,13,27,28^

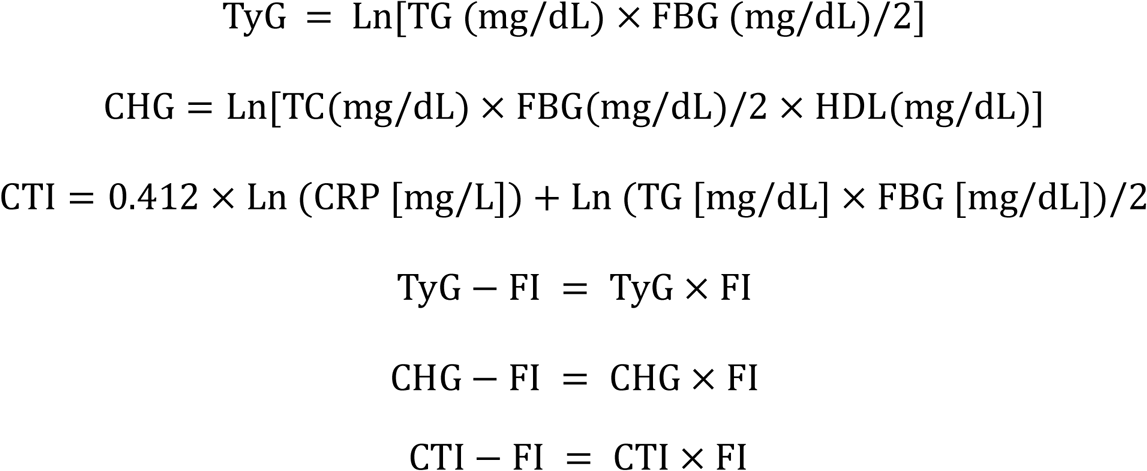

Men: eCRF (METs) = 21.2870 + (age×0.1654) – (age^2^×0.0023) – (BMI×0.2318) – (WC × 0.0337) – (rHR × 0.0390) + (moderate-to-vigorous PA × 0.6351) – (smoking×0.4263)

Women: eCRF (METs) = 14.7873 + (age× 0.1159) – (age^2^ × 0.0017) – (BMI× 0.1534) – (WC×0.0085) – (rHR×0.0364) + (moderate-to-vigorous PA×0.5987) – (smoking×0.2994)

### Endpoints

Mortality data from NHANES were obtained from the Public Use Linked Mortality File (cutoff December 31, 2019) via the unique identifier SEQN, with causes coded according to the International Statistical Classification of Diseases, 10th Revision. Mortality data in CHARLS were ascertained via biennial follow-up interviews up to 2020. Survival status was determined through interviews with family members or village/community committees. For deceased participants, the date of death was estimated as the midpoint between the last known alive wave and the wave when death was first reported. ACM included any death. CVM was defined as death from heart disease.

### Statistical analysis

Normality of continuous variables was assessed with the Shapiro-Wilk test. Normally distributed variables are presented as mean ± SD and were compared among three groups using one-way ANOVA. Non-normally distributed variables are reported as median [interquartile range (IQR)] with the Kruskal-Wallis test. Categorical variables are presented as counts (percentages), and the χ² test was used to compare differences between groups. Based on the medians of CTI-FI and eCRF, the participants were divided into three subgroups: (Low CTI-FI + High eCRF group, Consistent group, High CTI-FI + Low eCRF group).

After excluding variables with a variance inflation factor (VIF) >5 to control for multicollinearity, we performed LASSO regression with 10-fold cross-validation and selected lambda.1se to retain predictors with non-zero coefficients. We then included these variables in a multivariable Cox model to assess their predictive value. Meanwhile, a random forest model was run to rank the importance of all candidate variables. Weighted Cox proportional hazards models (for NHANES) and standard Cox proportional hazards models (for CHARLS) were used to evaluate the independent association of CTI-FI and eCRF with mortality outcomes in each cohort. CTI-FI was entered into the model as a continuous variable per 1-unit increase, and eCRF was entered per 1-MET (metabolic equivalent of task) increase. Four progressive adjustment models were constructed: unadjusted model; model 1 adjusted for age and sex; model 2 further adjusted for marital status and smoking; Model 3 ⅰ additionally adjusted for pulse pressure (PP), eGFR, and eCRF; Model 3 ⅱ additionally adjusted for PP, eGFR, and CTI-FI; Model 3 ⅲ additionally adjusted for PP and eGFR. RCS were fitted to model dose-response relationships of eCRF and CTI-FI with mortality risk separately for each sex, with knots placed at the 5th, 35th, 65th, and 95th percentiles of each variable. Kaplan-Meier survival curves were plotted, and the log-rank test was performed. Mediation and mutual mediation analyses between CTI-FI and eCRF, stratified by population (US vs. Chinese) and by sex, were performed to explore the potential pathways through which eCRF affects mortality outcomes and the bidirectional relationships between CTI-FI and eCRF.

For subgroup analyses, we stratified by age (45–59 vs. ≥60 years), sex, marital status, education level, hypertension, diabetes, and CKM stages (categorised as early/preventive [stages 0-2] vs. subclinical/high-risk [stage 3]). Interaction terms were included in the multivariable model to test for effect modification. Sensitivity analyses were performed to test the robustness of the findings, including complete-case analysis, exclusion of follow-up <2 years, landmark analyses with follow-up ≥5, 7, and 9 years, a two-step variable selection (LASSO lambda. min followed by Cox regression P < 0.05), external validation of all preceding steps using the CHARLS dataset, and Fine- Gray subdistribution hazard models for CVM to account for competing risk of non-CVM. E-values were calculated to evaluate potential confounding factors.

For ACM, the NHANES dataset was randomly split 70/30 into a training set (N = 4,666) and a test set (N = 1,996). For CVM, due to the limited number of events, internal validation was performed using 10-fold cross-validation to ensure stable performance estimates. Model discrimination and calibration were evaluated using time-dependent area under the receiver operating characteristic curve (time-dependent AUC), Harrell’s C-index, and calibration curves. Net reclassification improvement (NRI) and integrated discrimination improvement (IDI) were used to assess the added predictive value of eCRF, CHG-FI, TyG-FI, and CTI-FI beyond the basic model. A nomogram was constructed, and decision curve analysis (DCA) was performed to evaluate the clinical net benefit. Statistical analyses were performed using SPSS Statistics 26 and R software (version 4.5.2), with a two-sided P value < 0.05 considered statistically significant.

## Results

### Baseline data for study participants

**Table 1** presents baseline characteristics of the NHANES (n = 6,662) and CHARLS (n = 3,418) cohorts, overall and stratified into three subgroups: Low CTI-FI + High eCRF group (reference), Consistent group, and High CTI-FI + Low eCRF group. In NHANES, age was 60.8 ± 10.6 years, CTI-FI was 1.18 (IQR 0.76–1.79), and eCRF was 8.57 (IQR 7.10–10.26) metabolic equivalents (METs). In CHARLS, age was 58.9 ± 9.3 years, CTI-FI was 1.08 (IQR 0.61–1.83), and eCRF was 10.1 (IQR 8.97–11.96) METs.

**Table 1.** General baseline characteristics of the included population.

| Characteristic | Total | Low CTI-FI + High eCRF | Consistent group | High CTI-FI + Low eCRF | *P* value |
| --- | --- | --- | --- | --- | --- |
| NHANES cohort | (n = 6,662) | group (n = 2,079) | (n = 2,508) | group (n = 2,075) |  |
| Age (years) | 60.8 ± 10.6 | 56.2 ± 8.7 | 61.1 ± 10.3 | 65.2 ± 10.9 | <0.001 |
| Sex, male | 3242 (48.7%) | 1505 (72.4%) | 1224 (48.8%) | 513 (24.7%) | <0.001 |
| Marital status |  |  |  |  |  |
| Married or cohabiting | 4388 (65.9%) | 1597 (76.8%) | 1633 (65.1%) | 1158 (55.8%) | <0.001 |
| Divorced or others | 2274 (34.1%) | 482 (23.2%) | 875 (34.9%) | 917 (44.2%) |  |
| Education level |  |  |  |  |  |
| Below high school | 2035 (30.5%) | 505 (24.3%) | 737 (29.4%) | 793 (38.2%) | <0.001 |
| High school or above | 4627 (69.5%) | 1574 (75.7%) | 1771 (70.6%) | 1282 (61.8%) |  |
| Diabetes | 1084 (16.3%) | 80 (3.8%) | 356 (14.2%) | 648 (31.2%) | <0.001 |
| Hypertension | 3570 (53.6%) | 738 (35.5%) | 1321 (52.7%) | 1511 (72.8%) | <0.001 |
| Chronic kidney disease | 175 (2.6%) | 7 (0.3%) | 63 (2.5%) | 105 (5.1%) |  |
| Smoking status | 3338 (50.1%) | 1010 (48.6%) | 1324 (52.8%) | 1004 (48.4%) | 0.003 |
| Alcohol consumption | 4817 (72.3%) | 1696 (81.6%) | 1840 (73.4%) | 1281 (61.7%) | <0.001 |
| Physical activity | 1980 (29.7%) | 243 (11.7%) | 721 (28.7%) | 1016 (49%) | <0.001 |
| antihypertensive drug | 362 (5.4%) | 23 (1.1%) | 101 (4%) | 238 (11.5%) | <0.001 |
| antidiabetic drug | 3088 (46.4%) | 1220 (58.7%) | 1180 (47%) | 688 (33.2%) | <0.001 |
| CKM stages 0-2 | 5028 (75.5%) | 1788 (86%) | 1888 (75.3%) | 1352 (65.2%) | <0.001 |
| CKM stage 3 | 1634 (24.5%) | 291 (14%) | 620 (24.7%) | 723 (34.8%) | <0.001 |
| ACM | 1276 (19.2%) | 213 (10.2%) | 492 (19.6%) | 571 (27.5%) | <0.001 |
| CVM | 280 (4.2%) | 35 (1.7%) | 103 (4.1%) | 142 (6.8%) | <0.001 |
| Systolic blood pressure (mmHg) | 127 (116 - 140) | 123 (113 - 134) | 127 (117 - 141) | 131 (120 - 145) | <0.001 |
| Diastolic blood pressure (mmHg) | 72 (65 - 79) | 73 (67 - 80) | 72 (65 - 79) | 70 (62 - 77) | <0.001 |
| Pulse pressure (mmHg) | 54 (44 - 68) | 49 (41 - 59) | 55 (45 - 68) | 61 (49 - 76) | <0.001 |
| Fasting blood glucose (mg/dL) | 103 (96 - 115) | 100 (94 - 107) | 103 (96 - 114) | 109 (99 - 130) | <0.001 |
| HbA1c (%) | 5.6 (5.4 - 6.0) | 5.5 (5.2 - 5.7) | 5.6 (5.4 - 5.9) | 5.9 (5.5 - 6.5) | <0.001 |
| Tryglicerides (mg/dL) | 115 (82 - 168) | 102 (72 - 149) | 115 (83 - 169) | 131 (92 - 186) | <0.001 |
| Total cholesterol (mg/dL) | 202 (176 - 229) | 204 (181 - 230) | 202 (177 - 232) | 198 (171 - 226) | 0.001 |
| LDL-cholesterol (mg/dL) | 120 (98 - 144) | 125 (104 - 148) | 121 (98 - 145) | 114 (91 - 139) | <0.001 |
| HDL-cholesterol (mg/dL) | 52 (43 - 64) | 53 (44 - 65) | 52 (43 - 65) | 52 (42 - 63) | 0.001 |
| Body mass index (kg/m<sup>2</sup>) | 28.0 (24.8 - 32.0) | 25.7 (23.3 - 28.5) | 27.9 (25.0 - 31.4) | 31.7 (27.8 - 36.4) | <0.001 |
| Waist circumference (cm) | 99.2 (90.5 - 109.0) | 93.7 (85.4 - 101.4) | 98.6 (90.2 - 107.5) | 107.5 (97.7 - 117.6) | <0.001 |
| Heart rate | 68 (62 - 76) | 66 (60 - 72) | 69 (62 - 76) | 72 (64 - 80) | <0.001 |
| Blood urea nitrogen (mg/dL) | 14 (11 - 17) | 14 (11 - 16) | 14 (11 - 17) | 14 (11 - 18) | <0.001 |
| Uric acid | 5.4 (4.5 - 6.4) | 5.4 (4.5 - 6.4) | 5.4 (4.5 - 6.3) | 5.5 (4.6 - 6.5) | 0.001 |
| eGFR (mL/min/1.73m<sup>2</sup>) | 91 (76 - 101) | 94 (81 - 105) | 91 (77 - 101) | 85 (68 - 98) | <0.001 |
| C-reactive protein | 2.2 (1.0 - 4.7) | 1.3 (0.6 - 2.6) | 2.3 (1.0 - 4.5) | 3.6 (1.7 - 7.6) | <0.001 |
| Frailty index (FI) | 0.13 (0.09 - 0.20) | 0.08 (0.06 - 0.10) | 0.13 (0.09 - 0.18) | 0.20 (0.16 - 0.26) | <0.001 |
| TyG | 8.73 (8.34 - 9.16) | 8.56 (8.18 - 8.95) | 8.72 (8.36 - 9.15) | 8.94 (8.54 - 9.35) | <0.001 |
| CHG | 5.31 (5.05 - 5.59) | 5.25 (5.01 - 5.51) | 5.30 (5.04 - 5.58) | 5.37 (5.11 - 5.65) | <0.001 |
| CTI | 9.10 (8.51 - 9.66) | 8.70 (8.15 - 9.24) | 9.10 (8.54 - 9.64) | 9.47 (8.96 - 10.00) | <0.001 |
| TyG-FI | 1.14 (0.74 - 1.72) | 0.70 (0.48 - 0.91) | 1.14 (0.79 - 1.59) | 1.81 (1.42 - 2.35) | <0.001 |
| CHG-FI | 0.70 (0.45 - 1.04) | 0.43 (0.30 - 0.56) | 0.70 (0.48 - 0.98) | 1.10 (0.86 - 1.42) | <0.001 |
| CTI-FI | 1.18 (0.76 - 1.79) | 0.70 (0.48 - 0.92) | 1.18 (0.83 - 1.64) | 1.90 (1.50 - 2.50) | <0.001 |
| eCRF | 8.57 (7.10 - 10.26) | 10.4 (9.46 - 11.65) | 8.57 (7.50 - 9.97) | 6.84 (5.85 - 7.69) | <0.001 |
| Characteristic | Total | Low CTI-FI + High eCRF | Consistent group | High CTI-FI + Low eCRF | *P* value |
| CHARLS cohort | (n = 3,418) | group (n = 997) | (n = 1,425) | group (n = 996) |  |
| Age (years) | 58.9 ± 9.3 | 56.0 ± 7.9 | 58.2 ± 8.8 | 63.0 ± 9.8 | <0.001 |
| Sex, male | 1589 (46.5%) | 820 (82.2%) | 648 (45.5%) | 121 (12.1%) | <0.001 |
| Marital status |  |  |  |  |  |
| Married or cohabiting | 2872 (84%) | 897 (90%) | 1218 (85.5%) | 757 (76%) | <0.001 |
| Divorced or others | 546 (16%) | 100 (10%) | 207 (14.5%) | 239 (24%) |  |
| Education level |  |  |  |  |  |
| Below high school | 3084 (90.2%) | 825 (82.7%) | 1300 (91.2%) | 959 (96.3%) | <0.001 |
| High school or above | 334 (9.8%) | 172 (17.3%) | 125 (8.8%) | 37 (3.7%) |  |
| Diabetes | 152 (4.4%) | 10 (1%) | 52 (3.6%) | 90 (9%) | <0.001 |
| Hypertension | 704 (20.6%) | 97 (9.7%) | 280 (19.6%) | 327 (32.8%) | <0.001 |
| Chronic kidney disease | 198 (5.8%) | 50 (5%) | 80 (5.6%) | 68 (6.8%) | 0.208 |
| Smoking status | 1291 (37.8%) | 604 (60.6%) | 538 (37.8%) | 149 (15%) | 0.003 |
| Alcohol consumption | 1085 (31.7%) | 308 (30.9%) | 460 (32.3%) | 317 (31.8%) | 0.769 |
| Physical activity | 2362 (69.1%) | 812 (81.4%) | 980 (68.8%) | 570 (57.2%) | <0.001 |
| antihypertensive drug | 557 (16.3%) | 74 (7.4%) | 213 (14.9%) | 270 (27.1%) | <0.001 |
| antidiabetic drug | 105 (3.1%) | 8 (0.8%) | 35 (2.5%) | 62 (6.2%) | <0.001 |
| CKM stages 0-2 | 2570 (75.2%) | 794 (79.6%) | 1104 (77.5%) | 672 (67.5%) | <0.001 |
| CKM stage 3 | 848 (24.8%) | 203 (20.4%) | 321 (22.5%) | 324 (32.5%) | <0.001 |
| ACM | 391 (11.4%) | 74 (7.4%) | 170 (11.9%) | 147 (14.8%) | <0.001 |
| Systolic blood pressure (mmHg) | 127 (114 - 142) | 124 (113 - 136) | 127 (114 - 141) | 132 (117 - 149) | 0.004 |
| Diastolic blood pressure (mmHg) | 75 (67 - 83) | 74 (67 - 82) | 75 (67 - 84) | 75 (67 - 84) | 0.047 |
| Pulse pressure (mmHg) | 51 (44 - 61) | 50 (43 - 57) | 50 (44 - 60) | 56 (46 - 68) | 0.021 |
| Fasting blood glucose (mg/dL) | 102 (94 - 113) | 100 (92 - 109) | 102 (95 - 112) | 104 (96 - 117) | <0.001 |
| HbA1c (%) | 5.1 (4.9 - 5.4) | 5.1 (4.8 - 5.3) | 5.1 (4.8 - 5.4) | 5.2 (4.9 - 5.5) | 0.010 |
| Tryglicerides (mg/dL) | 104 (74 - 150) | 89 (66 - 132) | 104 (74 - 147) | 119 (85 - 167) | <0.001 |
| Total cholesterol (mg/dL) | 191 (167 - 215) | 186 (161 - 208) | 190 (168 - 214) | 198 (174 - 223) | 0.001 |
| LDL-cholesterol (mg/dL) | 114 (94 - 137) | 109 (91 - 132) | 115 (94 - 136) | 120 (98 - 143) | 0.003 |
| HDL-cholesterol (mg/dL) | 50 (41 - 60) | 50 (41 - 62) | 50 (41 - 60) | 48 (40 - 59) | 0.300 |
| Body mass index (kg/m²) | 23.1 (20.9 - 25.7) | 22.2 (20.4 - 24.3) | 23.1 (20.8 - 25.8) | 24.2 (21.9 - 27.0) | <0.001 |
| Waist circumference (cm) | 84.4 (77.5 - 91.7) | 82.0 (76.0 - 88.5) | 83.8 (77.0 - 91.0) | 88.1 (81.0 - 95.0) | <0.001 |
| Heart rate | 72 (65 - 78) | 70 (63 - 76) | 71 (65 - 78) | 74 (67 - 80) | <0.001 |
| Blood urea nitrogen (mg/dL) | 15 (13 - 18) | 15 (13 - 18) | 15 (12 - 18) | 15 (13 - 18) | 0.032 |
| Uric Acid | 4.3 (3.6 - 5.1) | 4.6 (3.8 - 5.5) | 4.2 (3.5 - 5.1) | 4.1 (3.4 - 4.9) | 0.001 |
| eGFR (mL/min/1.73m²) | 99 (89 - 105) | 101 (92 - 107) | 100 (90 - 106) | 95 (82 - 102) | 0.001 |
| C-reactive protein | 1.0 (0.5 - 2.0) | 0.9 (0.5 - 1.7) | 1.0 (0.5 - 1.0) | 1.2 (0.6 - 2.6) | 0.001 |
| Frailty index | 0.13 (0.07 - 0.21) | 0.07 (0.04 - 0.09) | 0.13 (0.08 - 0.21) | 0.21 (0.17 - 0.30) | <0.001 |
| TyG | 8.58 (8.20 - 9.01) | 8.40 (8.06 - 8.82) | 8.58 (8.22 - 8.99) | 8.74 (8.38 - 9.19) | <0.001 |
| CHG | 5.27 (5.02 - 5.56) | 5.19 (4.95 - 5.45) | 5.26 (5.03 - 5.55) | 5.37 (5.11 - 5.65) | <0.001 |
| CTI | 8.64 (8.12 - 9.21) | 8.39 (7.91 - 8.98) | 8.63 (8.16 - 9.19) | 8.88 (8.39 - 9.53) | <0.001 |
| TyG-FI | 1.07 (0.62 - 1.81) | 0.60 (0.34 - 0.78) | 1.07 (0.65 - 1.74) | 1.88 (1.42 - 2.62) | <0.001 |
| CHG-FI | 0.66 (0.38 - 1.12) | 0.36 (0.21 - 0.48) | 0.66 (0.39 - 1.04) | 1.16 (0.87 - 1.60) | <0.001 |
| CTI-FI | 1.08 (0.61 - 1.83) | 0.58 (0.34 - 0.79) | 1.08 (0.64 - 1.74) | 1.91 (1.44 - 2.65) | <0.001 |
| eCRF | 10.1 (8.97 - 11.96) | 12.17 (10.90 - 13.03) | 10.11 (9.16 - 11.73) | 8.80 (7.97 - 9.41) | <0.001 |

Data are presented as mean (standard deviation) or median (interquartile) or n (%); Abbreviations: CKM: cardiovascular-kidney-metabolic syndrome; ACM: all-cause mortality; CVM: cardiovascular mortality; HbA1c: hemoglobin A1c; LDL-cholesterol: low-density lipoprotein cholesterol; HDL-cholesterol: high-density lipoprotein cholesterol; eGFR: estimated glomerular filtration rate; TyG: triglyceride-glucose index; CHG: cholesterol, high-density lipoprotein, and glucose index; CTI: C-reactive protein-triglyceride-glucose index; TyG-FI: triglyceride glucose-frailty index; CHG-FI: cholesterol, high-density lipoprotein, glucose and frailty index; CTI-FI: C-reactive protein-triglyceride glucose index and frailty index; eCRF: non-exercise estimated cardiorespiratory fitness.

Baseline characteristics differed significantly across subgroups for most variables (all P < 0.05 except for CKD [P = 0.208], alcohol consumption [P = 0.769], and HDL-C [P = 0.300] in CHARLS). Compared with the reference group, individuals in the High CTI-FI + Low eCRF group were older, had higher FBG, HbA1c, TG, BMI, WC, CRP, and FI, along with a higher prevalence of diabetes, hypertension, and CKD, as well as lower eGFR and PA. Regarding mortality, in NHANES, the overall ACM was 19.2% and CVM was 4.2%. ACM increased progressively from 10.2% in the reference group to 19.6% in the Consistent group and 27.5% in the High CTI-FI+Low eCRF group; corresponding CVM rates were 1.7%, 4.1%, and 6.8%, respectively. In CHARLS, the overall ACM was 11.4%, with a similar stepwise increase across the three groups (7.4%, 11.9%, and 14.8%).

### Covariate Selection

After excluding collinear covariates (VIF >5), 25 covariates were subjected to LASSO regression with 10-fold cross-validation (**Table S5**). We selected lambda. 1se to obtain a parsimonious model that retained only the most clinically relevant covariates. Using the eight predictors selected by lambda.1se (age, sex, marital status, smoking, PP, eGFR, eCRF and CTI-FI), we further performed weighted Cox regression and random forest importance ranking. All eight variables were independently associated with the outcome in weighted Cox regression (all P < 0.05) (**Table 2**). Random forest ranking confirmed their importance (**Figure. 2**).

**Figure 2.**
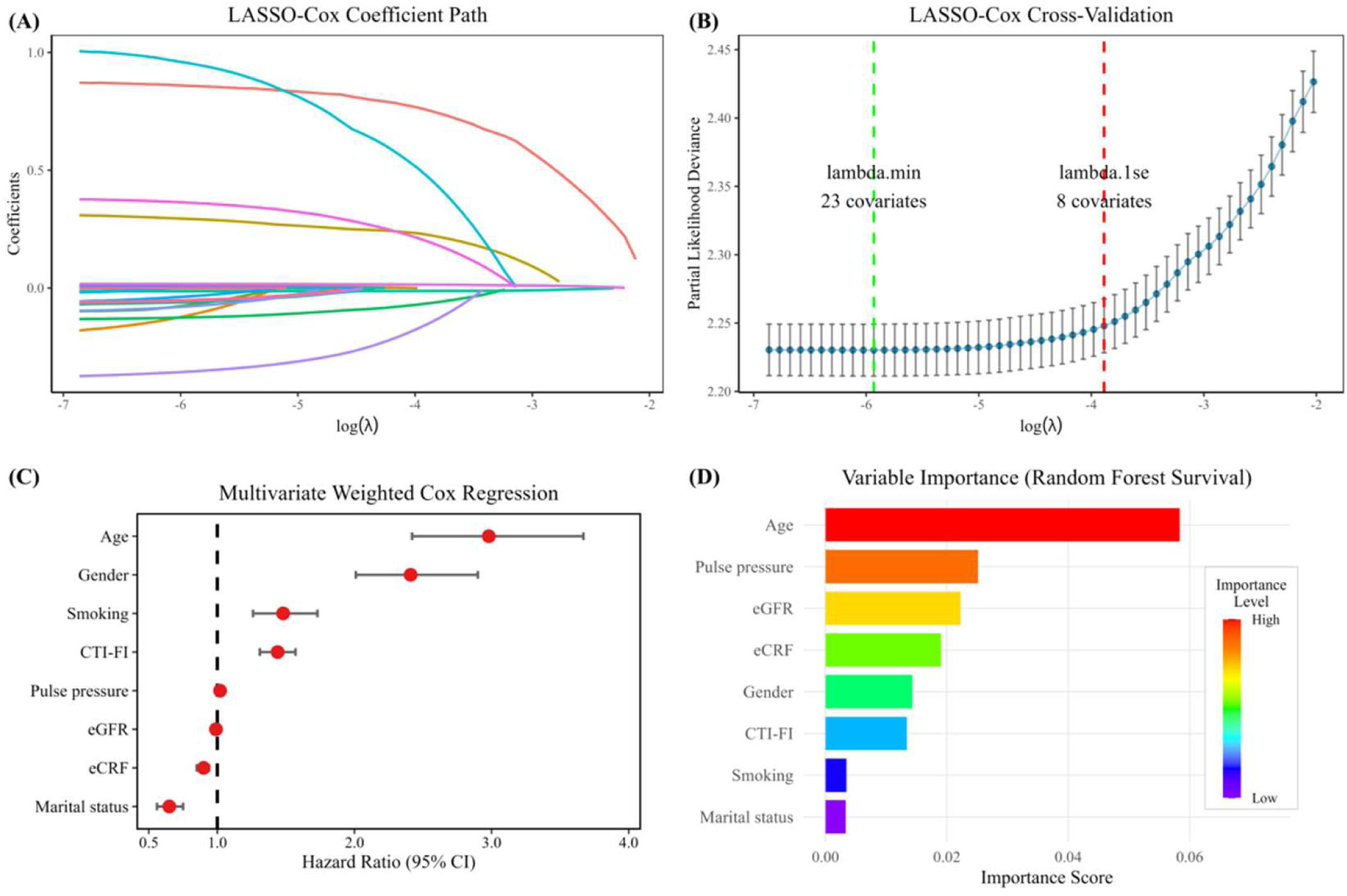
Covariate selection and variable importance. Legend: (A) LASSO-Cox regularization paths of regression coefficients. (B) 10-fold cross-validation for lambda selection; lambda. min (green) identifies 23 covariates, lambda. 1se (red) selects 8 robust covariates. (C) Weighted Cox regression forest plot of hazard ratios (HR, 95% CI) for all-cause mortality. (D) Random survival forest variable importance bar chart. Abbreviations: HR: hazard ratio; CI: confidence interval; CTI-FI, C-reactive protein-triglyceride glucose index and frailty index; eCRF, non-exercise estimated cardiorespiratory fitness; eGFR, estimated glomerular filtration rate.

**Table 2.** Weighted univariate and multivariate Cox regression analyses.

| Variable | All-cause mortality |  |  |  | Cardiovascular mortality |  |  |  |
| --- | --- | --- | --- | --- | --- | --- | --- | --- |
|  | Univariate analysis |  | Multivariate analysis |  | Univariate analysis |  | Multivariate analysis |  |
|  | HR (95% CI) | P value | HR (95% CI) | P value | HR (95% CI) | P value | HR (95% CI) | P value |
| Age | 5.44 (4.55 - 6.50) | <0.001 | 2.98 (2.42 - 3.67) | <0.001 | 6.29 (4.11 - 9.61) | <0.001 | 2.41 (1.52 - 3.84) | <0.001 |
| Gender | 1.19 (1.03 - 1.37) | 0.016 | 2.41 (2.01 - 2.90) | <0.001 | 1.31 (0.97 - 1.76) | 0.076 | 4.12 (2.89 - 5.87) | <0.001 |
| Marital status | 0.55 (0.47 - 0.64) | <0.001 | 0.65 (0.56 - 0.75) | <0.001 | 0.44 (0.32 - 0.60) | <0.001 | 0.53 (0.37 - 0.75) | <0.001 |
| Smoking | 1.54 (1.34 - 1.77) | <0.001 | 1.48 (1.26 - 1.73) | <0.001 | 1.24 (0.91 - 1.69) | 0.169 | 1.17 (0.84 - 1.64) | 0.347 |
| eGFR | 0.97 (0.97 - 0.97) | <0.001 | 0.99 (0.99 - 1.00) | 0.001 | 0.96 (0.95 - 0.97) | <0.001 | 0.99 (0.98 - 1.00) | 0.009 |
| Pulse pressure | 1.04 (1.03 - 1.04) | <0.001 | 1.02 (1.02 - 1.02) | <0.001 | 1.04 (1.04 - 1.05) | <0.001 | 1.03 (1.02 - 1.04) | <0.001 |
| CTI-FI | 1.62 (1.50 - 1.74) | <0.001 | 1.44 (1.31 - 1.57) | <0.001 | 1.81 (1.61 - 2.04) | <0.001 | 1.54 (1.33 - 1.79) | <0.001 |
| eCRF | 0.78 (0.75 - 0.82) | <0.001 | 0.90 (0.85 - 0.94) | <0.001 | 0.72 (0.67 - 0.79) | <0.001 | 0.82 (0.75 - 0.90) | <0.001 |

Abbreviations: HR: hazard ratio; CI: confidence interval; eGFR: estimated glomerular filtration rate; CTI-FI: C-reactive protein-triglyceride glucose index and frailty index; eCRF: estimated cardiorespiratory fitness. CTI-FI was entered into the model as a continuous variable per 1-unit increase, and eCRF was entered per 1-MET (metabolic equivalent of task) increase.

### CTI-FI, eCRF, and Their Joint Association with Mortality

For the NHANES cohort, CTI-FI achieved the greatest AUC values for predicting both ACM (0.605) and CVM (0.616) relative to all comparator metabolic-frailty indices. By contrast, eCRF (a protective factor) yielded AUCs of 0.636 (ACM) and 0.660 (CVM) (**Figure. 3**). In fully adjusted Model 3, per-unit increment in continuous CTI-FI independently predicted elevated mortality risk for ACM (HR 1.44, 95%CI 1.31–1.57) and CVM (HR 1.54, 95%CI 1.33–1.79), while higher eCRF was linked to reduced mortality risk (ACM: HR 0.90, 95%CI 0.85–0.94; CVM: HR 0.82, 95%CI 0.75–0.90; all P < 0.001). Relative to the reference group (Low CTI-FI + High eCRF), participants with High CTI-FI + Low eCRF exhibited substantially higher risks of ACM (HR 2.74, 95%CI 2.20–3.40) and CVM (HR 5.04, 95%CI 3.02–8.40; both P < 0.001) (**Table 3**).

**Figure 3.**
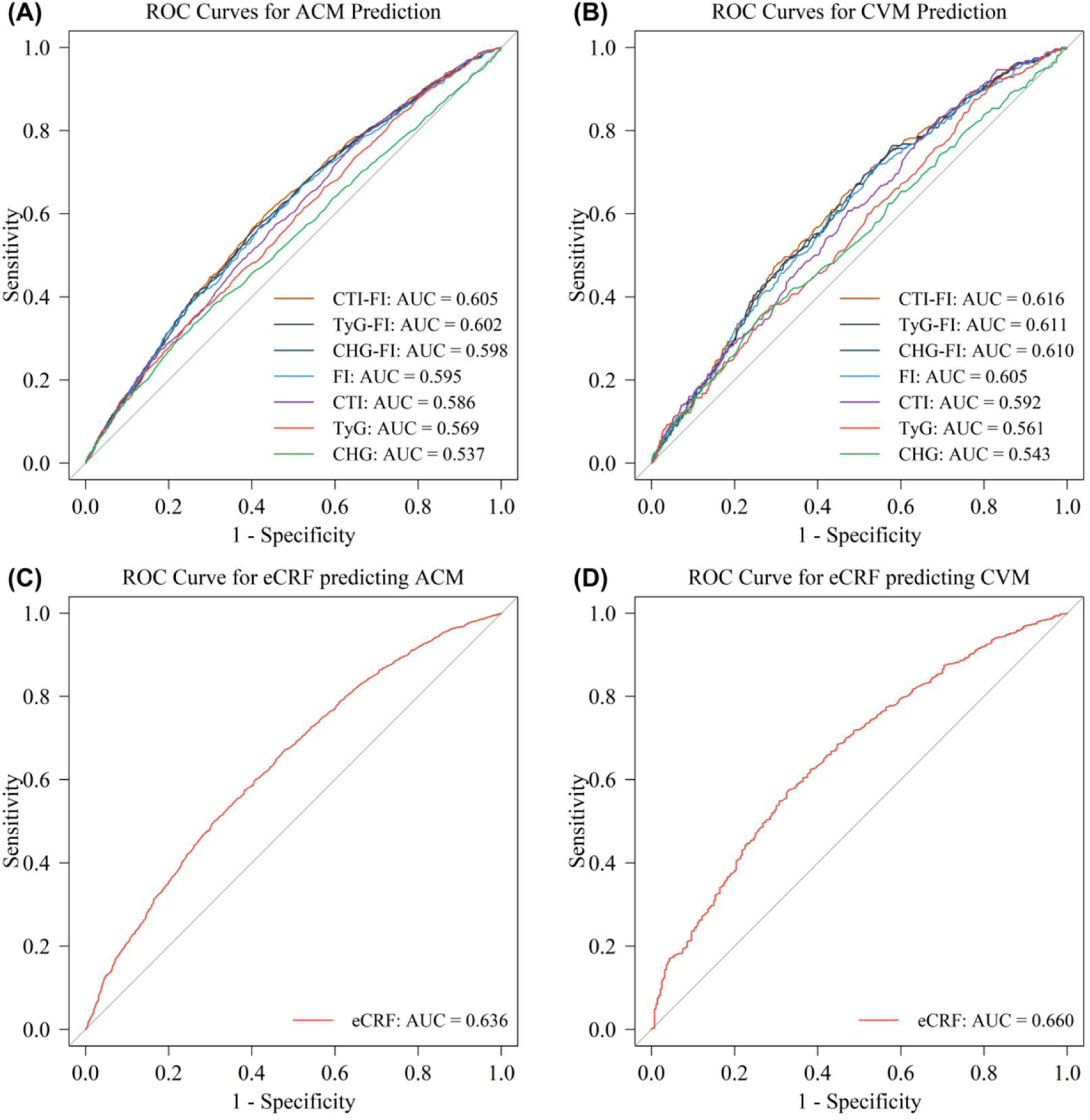
ROC analyses of CTI-FI and eCRF for mortality prediction. Legend: Time-independent ROC curves with area under the curve (AUC). CTI-FI versus other FI-derived and insulin-resistance indices for all-cause mortality (ACM) (A) and cardiovascular mortality (CVM) (B). eCRF for ACM (C) and CVM (D). Abbreviations: CTI-FI, C-reactive protein-triglyceride glucose index and frailty index; TyG-FI, triglyceride glucose-frailty index; CHG-FI, cholesterol, high-density lipoprotein, glucose and frailty index; FI, frailty index; CTI, C-reactive protein-triglyceride-glucose index; TyG, triglyceride-glucose index; CHG, cholesterol, high-density lipoprotein, and glucose index; eCRF, non-exercise estimated cardiorespiratory fitness.

**Table 3.** Association of CTI-FI and eCRF with mortality in NHANES.

| Association of CTI-FI and eCRF with all-cause mortality |  |  |  |  |  |  |  |  |
| --- | --- | --- | --- | --- | --- | --- | --- | --- |
| Variable | Unadjusted model |  | Model 1 |  | Model 2 |  | Model 3 |  |
|  | HR 95%CI | *P* value | HR 95%CI | *P* value | HR 95%CI | *P* value | HR 95%CI | *P* value |
| CTI-FI | 1.62 (1.50-1.74) | <0.001 | 1.63 (1.50-1.77) | <0.001 | 1.57 (1.44-1.71) | <0.001 | 1.44 (1.31-1.57) i | <0.001 |
| eCRF | 0.78 (0.75-0.82) | <0.001 | 0.81 (0.77-0.85) | <0.001 | 0.82 (0.78-0.86) | <0.001 | 0.90 (0.85-0.94) ii | <0.001 |
| Low CTI-FI +<br>High eCRF group | reference |  | reference |  | reference |  | reference iii |  |
| Consistent group | 2.25 (1.82-2.79) | <0.001 | 2.08 (1.68-2.58) | <0.001 | 1.96 (1.59-2.43) | <0.001 | 1.80 (1.46-2.22) | <0.001 |
| High CTI-FI +<br>Low eCRF group | 4.12 (3.31-5.11) | <0.001 | 3.65 (2.92-4.55) | <0.001 | 3.31 (2.66-4.13) | <0.001 | 2.74 (2.20-3.40) | <0.001 |
| *P* for trend | *P*<0.001 |  | *P*<0.001 |  | *P*<0.001 |  | *P*<0.001 |  |
| Association of CTI-FI and eCRF with cardiovascular mortality |  |  |  |  |  |  |  |  |
| Variable | Unadjusted model |  | Model 1 |  | Model 2 |  | Model 3 |  |
|  | HR 95%CI | *P* value | HR 95%CI | *P* value | HR 95%CI | *P* value | HR 95%CI | *P* value |
| CTI-FI | 1.81 (1.61-2.04) | <0.001 | 1.84 (1.62-2.09) | <0.001 | 1.76 (1.54-2.02) | <0.001 | 1.54 (1.33-1.79) i | <0.001 |
| eCRF | 0.72 (0.67-0.79) | <0.001 | 0.73 (0.67-0.79) | <0.001 | 0.74 (0.68-0.80) | <0.001 | 0.82 (0.75-0.90) ii | <0.001 |
| Low CTI-FI +<br>High eCRF group | reference |  | reference |  | reference |  | reference iii |  |
| Consistent group | 3.35 (2.04-5.49) | <0.001 | 3.28 (1.98-5.44) | <0.001 | 3.20 (1.93-5.31) | <0.001 | 2.81 (1.69-4.68) | <0.001 |
| High CTI-FI +<br>Low eCRF group | 7.12 (4.38-11.59) | <0.001 | 7.18 (4.38-11.78) | <0.001 | 6.71 (4.13-10.90) | <0.001 | 5.04 (3.02-8.40) | <0.001 |
| *P* for trend | *P*<0.001 |  | *P*<0.001 |  | *P*<0.001 |  | *P*<0.001 |  |
| Association of CTI-FI and eCRF with all-cause mortality after excluding missing data |  |  |  |  |  |  |  |  |
| Variable | Unadjusted model |  | Model 1 |  | Model 2 |  | Model 3 |  |
|  | HR 95%CI | *P* value | HR 95%CI | *P* value | HR 95%CI | *P* value | HR 95%CI | *P* value |
| CTI-FI | 1.61 (1.48-1.75) | <0.001 | 1.65 (1.51-1.81) | <0.001 | 1.58 (1.44-1.74) | <0.001 | 1.46 (1.32-1.61) i | <0.001 |
| eCRF | 0.78 (0.74-0.83) | <0.001 | 0.81 (0.77-0.86) | <0.001 | 0.83 (0.78-0.87) | <0.001 | 0.90 (0.85-0.96) ii | <0.001 |
| Low CTI-FI +<br>High eCRF group | reference |  | reference |  | reference |  | reference iii |  |
| Consistent group | 2.31 (1.82-2.92) | <0.001 | 2.18 (1.73-2.74) | <0.001 | 2.04 (1.62-2.56) | <0.001 | 1.85 (1.47-2.34) | <0.001 |
| High CTI-FI +<br>Low eCRF group | 3.95 (3.08-5.06) | <0.001 | 3.60 (2.84-4.56) | <0.001 | 3.20 (2.54-4.04) | <0.001 | 2.63 (2.08-3.33) | <0.001 |
| *P* for trend | *P*<0.001 |  | *P*<0.001 |  | *P*<0.001 |  | *P*<0.001 |  |
| Association of CTI-FI and eCRF with cardiovascular mortality after excluding missing data |  |  |  |  |  |  |  |  |
| Variable | Unadjusted model |  | Model 1 |  | Model 2 |  | Model 3 |  |
|  | HR 95%CI | *P* value | HR 95%CI | *P* value | HR 95%CI | *P* value | HR 95%CI | *P* value |
| CTI-FI | 1.74 (1.54-1.97) | <0.001 | 1.81 (1.57-2.09) | <0.001 | 1.73 (1.49-2.00) | <0.001 | 1.50 (1.27-1.77) i | <0.001 |
| eCRF | 0.73 (0.67-0.80) | <0.001 | 0.73 (0.67-0.80) | <0.001 | 0.74 (0.67-0.80) | <0.001 | 0.81 (0.73-0.89) ii | <0.001 |
| Low CTI-FI +<br>High eCRF group | reference |  | reference |  | reference |  | reference iii |  |
| Consistent group | 3.75 (2.10-6.71) | <0.001 | 3.90 (2.17-7.03) | <0.001 | 3.78 (2.09-6.84) | <0.001 | 3.32 (1.82-6.06) | <0.001 |
| High CTI-FI +<br>Low eCRF group | 6.89 (4.01-11.86) | <0.001 | 7.69 (4.49-13.16) | <0.001 | 7.12 (4.14-12.23) | <0.001 | 5.45 (3.08-9.64) | <0.001 |
| *P* for trend | *P*<0.001 |  | *P*<0.001 |  | *P*<0.001 |  | *P*<0.001 |  |

Abbreviations: CTI-FI: C-reactive protein-triglyceride glucose index and frailty index; eCRF: non-exercise estimated cardiorespiratory fitness; HR: hazard ratio; CI: confidence interval
CTI-FI was entered into the model as a continuous variable per 1-unit increase, and eCRF was entered per 1-MET (metabolic equivalent of task) increase
Model 1 adjusted: age and sex
Model 2 adjusted: age, sex, marital status, and smoking
Model 3 i adjusted: age, sex, marital status, smoking, pulse pressure, estimated glomerular filtration rate, and eCRF
Model 3 ii adjusted: age, sex, marital status, smoking, pulse pressure, estimated glomerular filtration rate, and CTI-FI
Model 3 iii adjusted: age, sex, marital status, smoking, pulse pressure, estimated glomerular filtration rate

In the CHARLS cohort with ACM as the sole available endpoint, CTI-FI still yielded the highest discriminatory performance (AUC = 0.645) across all tested metabolic-frailty indices, with an eCRF AUC of 0.550 (**Figure. S1**). After full multivariable adjustment, continuous CTI-FI conferred excess ACM risk (HR 1.29, 95%CI 1.19– 1.40), and elevated eCRF was protective against ACM (HR 0.88, 95%CI 0.82–0.95; both P < 0.001). Consistently, the High CTI-FI + Low eCRF subgroup was associated with significantly greater ACM risk versus the reference profile (HR 2.27, 95%CI 1.62– 3.17; P < 0.001) (**Table S6**).

### Subgroup Analysis, Sensitivity Analysis, and E-values

Subgroup analyses revealed that nearly all interaction P-values for CTI-FI and eCRF across predefined covariates were >0.05, with the sole exception of the CTI-FI by gender subgroup (P for interaction <0.001), indicating that effect modification was not significant for the remaining covariates. Stratified by CKM classification, the adverse effect of CTI-FI and protective effect of eCRF remained significant in both CKM stage 0–2 and stage 3 for ACM and CVM in NHANES (**Figure. 4**).

**Figure 4.**
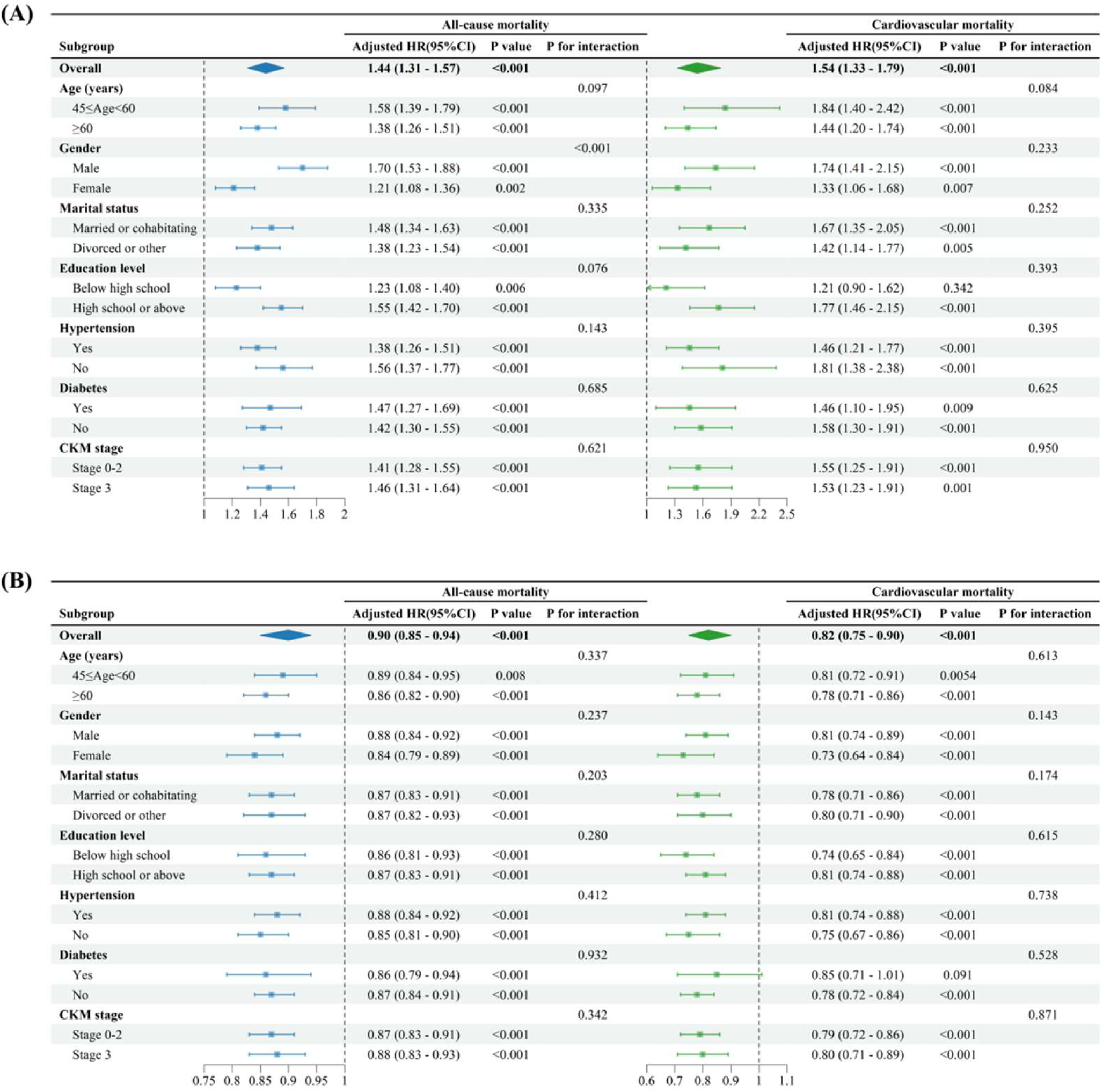
Subgroup analyses CTI-FI and eCRF with ACM and CVM. Legend: Forest plots of adjusted HR (95% CI) with interaction P-values. (A) CTI-FI stratified by age, sex, marital status, education level, hypertension, diabetes, and CKM stage. (B) eCRF using the same strata. Abbreviations: ACM, all-cause mortality; CVM, cardiovascular mortality; HR, hazard rate; CI, confidence interval; CTI-FI, C-reactive protein-triglyceride glucose index and frailty index; eCRF, non-exercise estimated cardiorespiratory fitness; CKM, cardiovascular-kidney-metabolic syndrome.

Sequential Cox models (from unadjusted to fully adjusted Model 3) confirmed stable risk estimates for CTI-FI, eCRF, and their combined grouping in both cohorts. Multiple prespecified sensitivity analyses (including follow-up restriction, landmark analysis, LASSO-based variable selection, and external validation) all yielded consistent HR magnitudes and significance (**Table 3 and Tables S6-11**). Similarly, Fine-Gray subdistribution hazard models treating non-CVM as a competing event confirmed the robustness of the associations for CVM (**Table S12**). E-values quantifying robustness against unmeasured confounding are summarized in **Table S13**. For ACM, E-values were 2.24 (CTI-FI per-unit HR = 1.44), 1.46 (eCRF per-unit HR = 0.90), and 4.92 (High CTI-FI + Low eCRF vs. Low CTI-FI + High eCRF). For CVM, corresponding E-values were 2.45, 1.74, and 9.55. All E-values exceeded 1.46, with the majority above 1.5, indicating moderate to strong robustness against potential unmeasured confounding.

### Restricted Cubic Spline Analysis

RCS analyses revealed distinct dose–response patterns for CTI-FI and eCRF (**Figure. 5** for NHANES; **Figure. S2** for CHARLS). In NHANES, CTI-FI showed a non-linear association with ACM (P for non-linearity < 0.05): below the inflection cutoff of 1.95, each unit increase in CTI-FI was associated with a 61% higher ACM risk (HR 1.61; 95% CI 1.42–1.83); above this threshold, the association attenuated (HR 1.18; 95% CI 1.06– 1.33). For CVM, CTI-FI followed a linear trend, with significant risk increase only above 1.95 (HR = 1.40). In contrast, eCRF exhibited linear inverse associations with both ACM and CVM, with protective effects confined below the median cutoff of 8.57 (ACM: HR = 0.87; CVM: HR = 0.80) and no significant association above this value (**Table 4**). In CHARLS, both CTI-FI and eCRF showed monotonic dose–response relationships, with higher CTI-FI associated with worse survival and higher eCRF with better survival.

**Figure 5.**
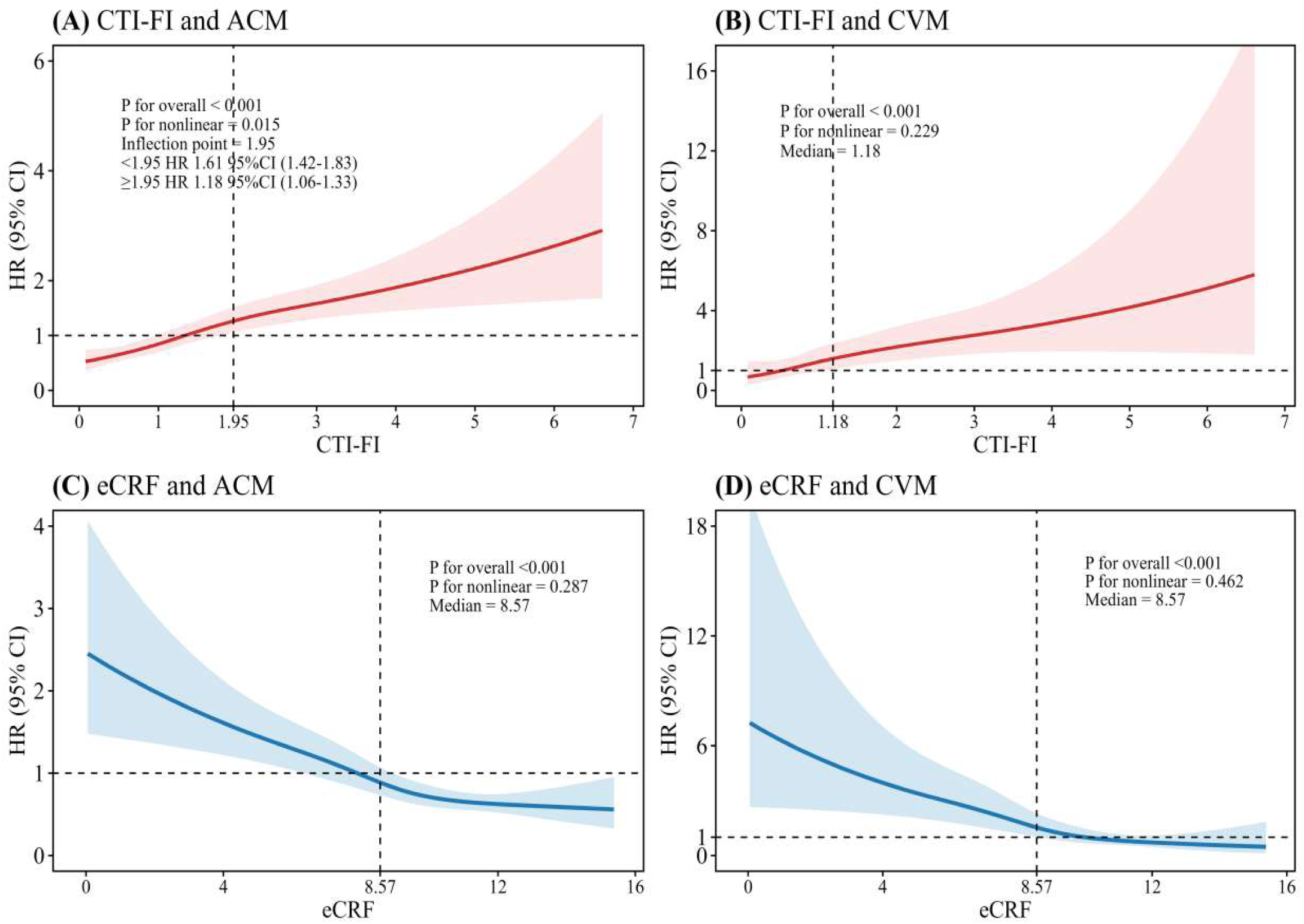
Restricted cubic spline (RCS) curves for CTI-FI and eCRF. Legend: Adjusted RCS plots with knots at 5th, 35th, 65th, and 95th percentiles. CTI-FI models adjusted for age, sex, marital status, smoking, pulse pressure, eGFR, and eCRF; eCRF models adjusted for the same covariates plus CTI-FI. (A) CTI-FI nonlinear with all-cause mortality (ACM). (B) CTI-FI linear with cardiovascular mortality (CVM). (C) eCRF nonlinear with ACM. (D) eCRF nonlinear with CVM. Shaded areas: 95% confidence interval of adjusted hazard rate. Abbreviations: CTI-FI, C-reactive protein-triglyceride glucose index and frailty index; eCRF, non-exercise estimated cardiorespiratory fitness.

**Table 4.**
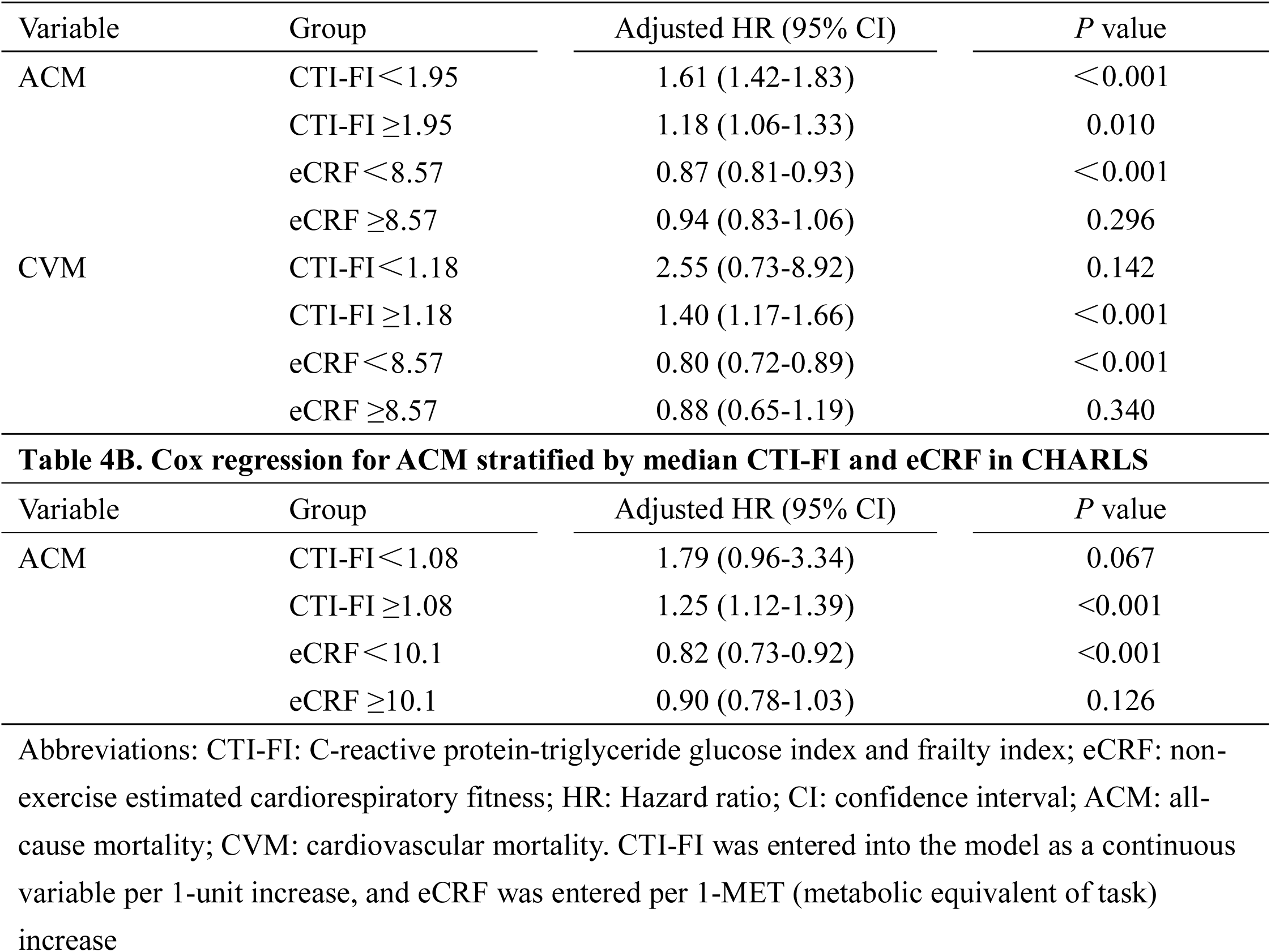
A. Cox regression for ACM stratified by CTI-FI inflection point and eCRF median in NHANES.

### Kaplan–Meier Survival Analysis

Kaplan–Meier curves stratified by median splits of individual biomarkers and the three composite phenotypes confirmed the dose–response findings (all log-rank P < 0.001; **Figure. S3** for NHANES, **Figure. S4** for CHARLS). In NHANES, participants with CTI-FI above the median had significantly worse ACM and CVM survival, whereas those with eCRF above the median had better long-term survival. When stratified by the combined grouping (reference: Low CTI-FI + High eCRF), survival declined sequentially from the reference to the Consistent group and then to the High CTI-FI + Low eCRF group. The same hierarchical survival gradient was reproduced in CHARLS for ACM, with the High CTI-FI + Low eCRF subgroup showing the poorest survival.

### Mediation Analysis for Reciprocal Pathways Between CTI-FI and eCRF

Counterfactual mediation analyses quantified bidirectional mediating effects between CTI-FI and eCRF on mortality (**Figure. 6**). In NHANES, eCRF mediated 16.4% of CTI-FI-associated ACM risk and 23.5% of CVM risk; reciprocally, CTI-FI mediated 32.2% (ACM) and 23.9% (CVM) of eCRF’s protective effect (all P < 0.001). Sex- stratified analyses revealed larger mediated fractions in females than in males (e.g., eCRF mediating CTI-FI-related CVM: 36.8% in females vs. 20.1% in males) (**Figure. S5**). In CHARLS, the bidirectional mediation patterns were replicated: eCRF mediated 6.8% of CTI-FI-associated ACM risk, and CTI-FI mediated 16.7% of eCRF’s protective effect (both P < 0.001). Sex-specific analyses in CHARLS showed that both pathways remained significant in females, whereas only the CTI-FI → eCRF pathway was significant in males (**Figure. S6**).

**Figure 6.**
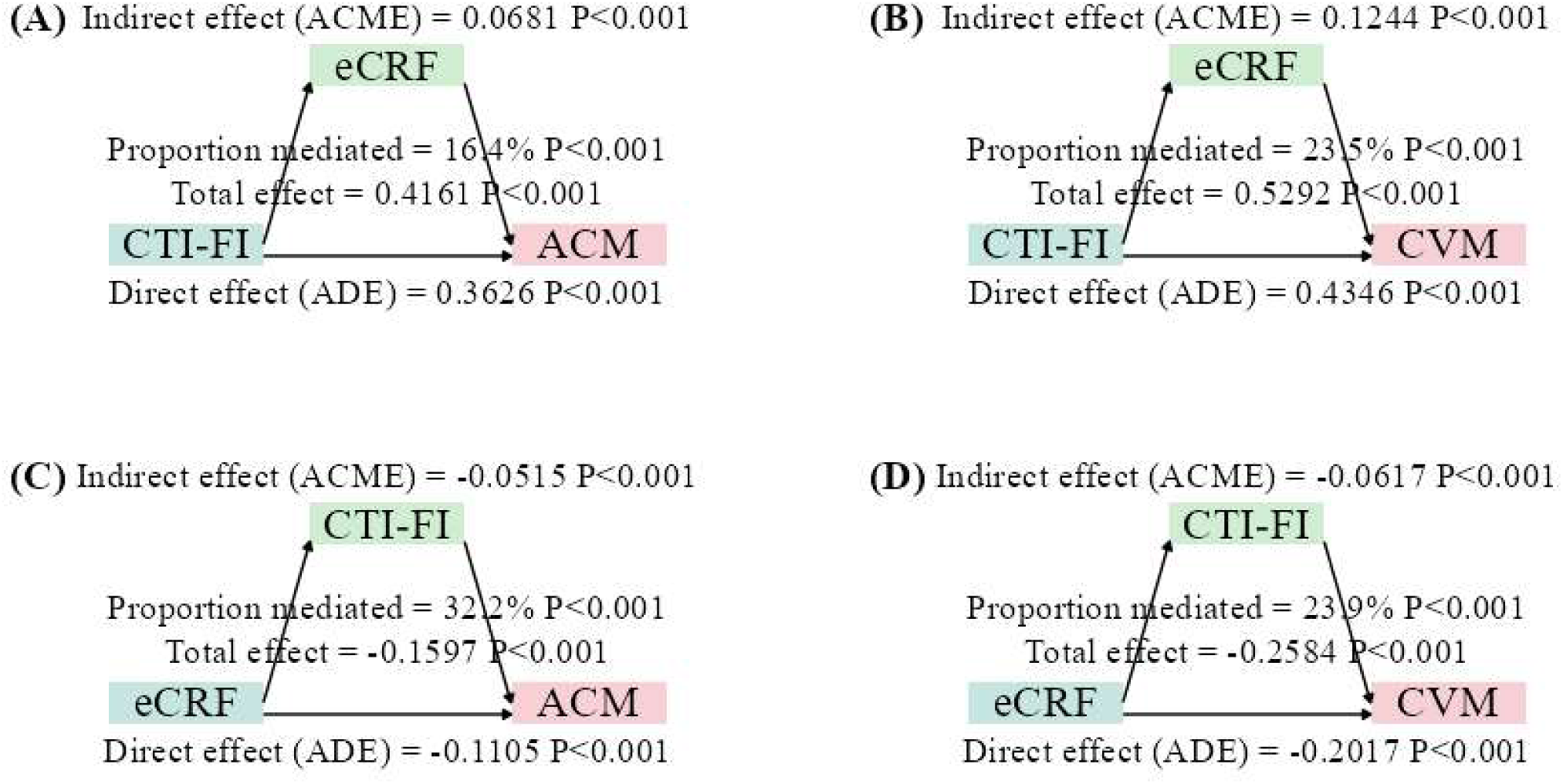
Bidirectional mediation among CTI-FI, eCRF, and mortality. Legend: Mediation analysis decomposing total effect into natural direct and average causal mediated effects. eCRF mediating CTI-FI to all-cause mortality (ACM) (A) and cardiovascular mortality (CVM) (B). CTI-FI mediating eCRF to ACM (C) and CVM (D). Abbreviations: CTI-FI, C-reactive protein-triglyceride glucose index and frailty index; eCRF, non-exercise estimated cardiorespiratory fitness; HR, hazard rate; CI, confidence interval.

### Incremental Predictive Value of Adding CTI-FI and eCRF

CTI-FI demonstrated slightly superior incremental performance compared to TyG-FI and CHG-FI, with numerically higher NRI values in several comparisons. Using a basic model including eCRF as the reference, the categorical NRI for CVM was 0.400 when CTI-FI was further added, versus 0.352 for TyG-FI and 0.338 for CHG-FI in NHANES. Similar patterns were observed in CHARLS (0.337, 0.314, and 0.309, respectively). Stepwise addition of eCRF alone and then with CTI-FI progressively improved model discrimination: in NHANES, baseline ACM AUC (0.765) increased to 0.773 with eCRF and to 0.777 with both biomarkers; for CVM, from 0.770 to 0.784 and 0.787, respectively. In CHARLS, AUC rose from 0.779 to 0.786 with eCRF and to 0.798 with CTI-FI+eCRF (**Table 5**). Time-dependent C-index values were stable across training (0.780), internal cross-validation (0.778), and external CHARLS validation (0.780); calibration curves closely followed the ideal diagonal (**Table S14, Figure. 7**).

**Figure 7.**
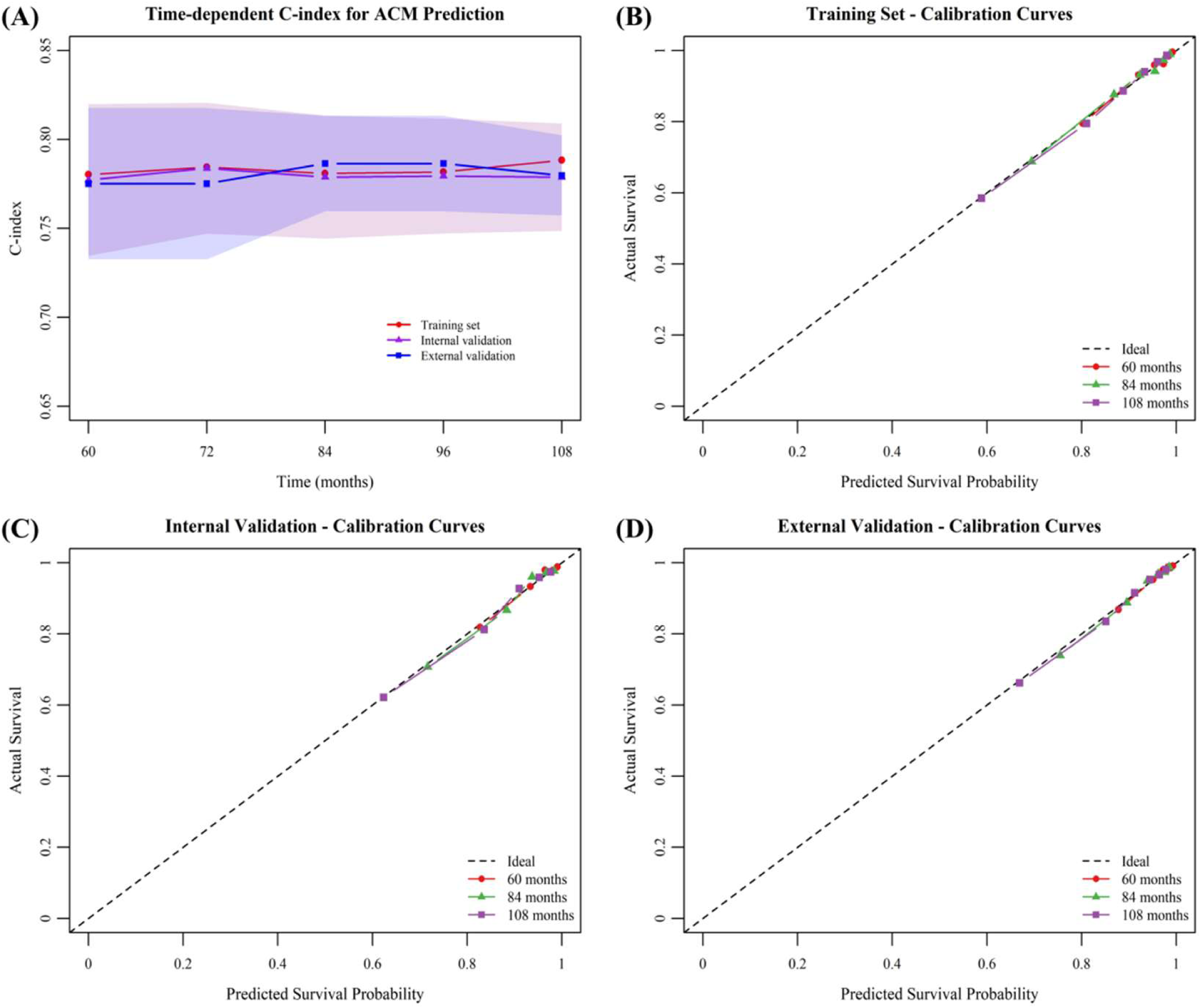
Model validation using time-dependent C-index and calibration curves Legend: (A) Time-dependent C-index (concordance index) in training, internal validation, and external validation cohorts. Calibration curves at 5, 7, and 9 years for training (B), internal validation (C), and external validation (D) cohorts; the diagonal dashed line indicates ideal calibration. Abbreviations: CTI-FI, C-reactive protein-triglyceride glucose index and frailty index; eCRF, non-exercise estimated cardiorespiratory fitness.

**Table 5.** Additional prognostic value of CTI-FI, eCRF, and other measures.

| Model | AUC (95%CI) | P-value | NRI (95%CI) | P-value | IDI (95%CI) | P-value |
| --- | --- | --- | --- | --- | --- | --- |
| **All-cause mortality (NHANES)** |  |  |  |  |  |  |
| Basic model | 0.765 (0.751-0.779) |  | Ref |  | Ref |  |
| +eCRF | 0.773 (0.759-0.786) | <0.001 | 0.237 (0.176-0.297) | <0.001 | 0.011 (0.008-0.014) | <0.001 |
| +eCRF+CHG-FI | 0.777 (0.763-0.790) | <0.001 | 0.291 (0.231-0.352) | <0.001 | 0.016 (0.012-0.019) | <0.001 |
| +eCRF+TyG-FI | 0.777 (0.763-0.790) | <0.001 | 0.286 (0.225-0.347) | <0.001 | 0.016 (0.012-0.020) | <0.001 |
| +eCRF+CTI-FI | 0.777 (0.763-0.791) | <0.001 | 0.283 (0.222-0.344) | <0.001 | 0.016 (0.013-0.020) | <0.001 |
| **Cardiovascular mortality (NHANES)** |  |  |  |  |  |  |
| Basic model | 0.770 (0.744-0.797) |  | Ref |  | Ref |  |
| +eCRF | 0.784 (0.759-0.810) | 0.007 | 0.384 (0.266-0.502) | <0.001 | 0.010 (0.006-0.014) | <0.001 |
| +eCRF+CHG-FI | 0.787 (0.761-0.812) | 0.003 | 0.373 (0.254-0.492) | <0.001 | 0.011 (0.007-0.016) | <0.001 |
| +eCRF+TyG-FI | 0.787 (0.762-0.812) | 0.002 | 0.382 (0.264-0.501) | <0.001 | 0.012 (0.007-0.016) | <0.001 |
| +eCRF+CTI-FI | 0.787 (0.762-0.812) | 0.002 | 0.400 (0.281-0.518) | <0.001 | 0.012 (0.007-0.017) | <0.001 |
| **All-cause mortality (CHARLS)** |  |  |  |  |  |  |
| Basic model | 0.779 (0.755-0.804) |  | Ref |  | Ref |  |
| +eCRF | 0.786 (0.761-0.810) | 0.028 | 0.147 (0.042-0.252) | 0.006 | 0.010 (0.005-0.015) | <0.001 |
| +eCRF+CHG-FI | 0.797 (0.773-0.821) | <0.001 | 0.319 (0.214-0.424) | <0.001 | 0.024 (0.015-0.032) | <0.001 |
| +eCRF+TyG-FI | 0.797 (0.774-0.821) | <0.001 | 0.319 (0.214-0.423) | <0.001 | 0.024 (0.015-0.033) | <0.001 |
| +eCRF+CTI-FI | 0.798 (0.774-0.822) | <0.001 | 0.337 (0.232-0.442) | <0.001 | 0.026 (0.017-0.035) | <0.001 |

Basic models were adjusted for age, sex, marital status, smoking, pulse pressure, and estimated glomerular filtration rate. Abbreviations: CTI-FI: C-reactive protein-triglyceride glucose index and frailty index; eCRF: non-exercise estimated cardiorespiratory fitness; AUC: area under the curve; NRI: net reclassification improvement; IDI: integrated discrimination improvement; CI: confidence interval; TyG-FI: Triglyceride-glucose and frailty indices; CHG-FI: cholesterol, high-density lipoprotein, glucose and frailty indices.

The model containing CTI-FI+eCRF showed a slightly higher overall C-index for CVM prediction (0.825) than the Framingham Risk Score (0.806) in 10-fold cross-validation, with consistently higher time-dependent C-index across 5–9 years (**Table S15, Figure. S7**). A nomogram integrating conventional covariates, eGFR, PP, eCRF, and CTI-FI was constructed to predict 5-, 7-, and 9-year ACM probability (**Figure. S8**), and DCA suggested higher net clinical benefit than the basic model across most threshold probabilities (**Figure. S9**).

## Discussion

In this cross-cohort analysis of US (NHANES) and Chinese (CHARLS) populations, we evaluated the predictive performance of a composite metabolic-frailty index, CTI-FI, alongside eCRF. First, CTI-FI consistently showed higher discriminatory ability for ACM and CVM than other metabolic-frailty indices (TyG-FI, CHG-FI, FI) in both cohorts. Second, CTI-FI was independently associated with increased mortality risk, whereas higher eCRF was independently protective. Third, individuals with the High CTI-FI + Low eCRF phenotype faced substantially elevated mortality compared with the Low CTI-FI + High eCRF reference, and adding both biomarkers to a basic risk model significantly improved predictive performance, achieving a higher C-index for CVM than the Framingham Risk Score. Fourth, bidirectional mediation analyses revealed that eCRF partially mediated the excess risk linked to CTI-FI, and vice versa, with larger mediated fractions in females. Finally, all key findings were replicated in the external CHARLS cohort, and decision curve analysis supported the clinical net benefit of the combined model. Together, these results position CTI-FI and eCRF as complementary, mechanistically interlinked predictors that refine mortality risk stratification across diverse populations.

In this study, CTI-FI consistently outperformed TyG-FI, CHG-FI, and the traditional FI in predicting both ACM and CVM. This superiority stems from CTI-FI’s integration of three domains: metabolic dysregulation, chronic inflammation, and cumulative deficits.^13^ Its core component, CRP—a marker of low-grade systemic inflammation— not only participates in IR and atherosclerosis but also directly promotes muscle catabolism and reduces exercise tolerance, thereby accelerating frailty.^29–33^ In contrast, TyG-FI reflects only glucolipid metabolism, CHG-FI captures only long-term glycemic control, and FI, despite covering multisystem deficits, lacks specific metabolic and inflammatory information.^10,12^ The advantage of CTI-FI lies in its organic combination of metabolic burden, inflammatory state, and physiological vulnerability, providing a more complete capture of the complex pathophysiological network driving mortality risk.^13^

Cut-off analysis further revealed clinically actionable thresholds. In NHANES, CTI-FI exhibited a non-linear association with ACM: below 1.95, each unit increase was associated with a significantly elevated risk; above this value, the risk gain attenuated markedly, suggesting a “ saturation effect ” . This finding aligns with the limited resilience of biological systems-beyond a certain critical point, the incremental risk from additional metabolic-inflammatory-frailty accumulation diminishes. Clinically, the threshold of 1.95 can be used to identify extremely high-risk individuals, and more importantly, it suggests that when CTI-FI is already markedly elevated, intervention efforts should shift from “further lowering the value” to “stabilizing the current state and preventing target-organ damage”. Moreover, CTI-FI showed a linear trend with CVM, with a significant risk increase, only above 1.18, indicating that cardiovascular endpoints are more sensitive to high levels of this index. Collectively, the superior discriminative ability of CTI-FI is rooted in its pathophysiological rationale and is supported by a clear dose-response pattern that facilitates clinical translation.

Beyond CTI-FI, however, the potential protective role of eCRF within the CKM framework remains to be elucidated. We found that eCRF maintained significant and similarly sized protective effects across both CKM stages 0–2 and stage 3, with HRs for ACM of approximately 0.87–0.88 and for CVM of approximately 0.79–0.80. These findings are broadly consistent with prior studies that have reported an inverse association between eCRF and mortality in other populations.^18–20,23^ However, none of those studies specifically focused on CKM syndrome. Notably, the recent work evaluated the prognostic value of eCRF in patients with CKD – a condition that shares substantial pathophysiological overlap with advanced CKM syndrome.^20,21^ The consistency of effect sizes between our CKM subgroups and the CKD population suggests that the survival benefit of improving eCRF is robust across different stages of cardiorenal-metabolic disease. Taken together, these findings indicate that whether patients are in early metabolic risk or have already developed cardiovascular or kidney disease, enhancing eCRF confers comparable survival benefits, and the CKM stage does not modify the direction or magnitude of eCRF’s protective effect.

Having established the distinct roles of CTI-FI and eCRF, we next integrated them and found that the combination of high CTI-FI and low eCRF identified a particularly high-risk phenotype. In this study, compared with the reference group, the High CTI-FI+Low eCRF group had approximately a 2.3- to 2.7-fold higher ACM risk and a 5-fold higher CVM risk, far exceeding the sum of the effects of each indicator, suggesting substantial multiplicative interaction. This synergistic amplification may occur through a vicious cycle. On one hand, CTI-FI, as a composite biomarker integrating CRP and the TyG index, captures the cumulative burden of both IR and systemic inflammation.^13,31,34,35^ Mechanistically, TyG index— another core component of CTI-FI— is significantly associated with sarcopenia, with oxidative stress and chronic inflammation acting as key mediators.^36–38^ Hyperglycemia and IR impair mitochondrial function through energy imbalance, oxidative stress damage, and abnormalities in mitochondrial dynamics, thereby compromising muscle mass, strength, and eCRF.^39–41^ On the other hand, low eCRF is inversely associated with resting inflammatory markers, and each 1-MET increase in CRF reduces ACM by 14% and CVM by 16%.^23^ A large prospective study further demonstrated that high eCRF is associated with slower accumulation of chronic diseases and delayed multimorbidity onset.^19,28^ Additionally, exercise training has been shown to reduce systemic inflammatory markers and improve IR.^41,42^ Conversely, IR promotes lipotoxicity, adipose tissue dysfunction, and inflammatory cascades, which further impair insulin receptor function.^43,44^ Thus, CTI-FI and eCRF reinforce each other bidirectionally: a high metabolic-inflammatory burden drives eCRF decline, whereas low eCRF in turn exacerbates metabolic inflammation, forming a vicious cycle that accelerates cardiovascular events and mortality. Based on this rationale, we incorporated both CTI-FI and eCRF into a new clinical prediction model. In our validation, the model achieved a C-index of 0.78 for ACM and 0.83 for CVM, with the latter surpassing 0.81 obtained by the Framingham Risk Score (FRS). Furthermore, DCA demonstrated that the new model provided greater net clinical benefit than the basic model. These findings collectively confirm the superior value of combining CTI-FI and eCRF.

To elucidate the directional nature of these associations, mediation analyses revealed a bidirectional causal relationship. In NHANES, eCRF mediated a significant proportion of CTI-FI-associated mortality risk, and conversely, CTI-FI mediated a substantial portion of eCRF’s protective effect. The proportions were asymmetric, suggesting that the protection conferred by eCRF via lowering CTI-FI was somewhat stronger than the risk mediated by CTI-FI via suppressing eCRF. This hints that improving eCRF may be more effective in “ cutting off ” the adverse pathway than simply controlling metabolic-inflammatory burden.

Sex differences were another key finding. Across all pathways, the mediated proportions were larger in women than in men (for example, the proportion of CTI-FI-related CVM risk mediated by eCRF was approximately 1.8-fold higher in women). Potential biological explanations include the following. Estrogen possesses anti-inflammatory, antioxidant, and pro-angiogenic properties that may enhance eCRF’s capacity to buffer metabolic inflammation.^45^ However, as our study focuses on middle-aged and older adults, the sharp decline in estrogen after menopause substantially attenuates its protective effects, which may render the mediation pathway more prominent. Regarding body composition and fat distribution, postmenopausal women exhibit increased visceral adiposity that exacerbates IR and systemic inflammation.^46,47^ Moreover, the inverse association between low eCRF and both inflammation and fat accumulation is stronger in women, thereby amplifying the mediation effect.^48^ With respect to eCRF modulation of inflammation, previous studies indicate that exercise-induced reductions in CRP are more pronounced in women; consequently, improvements in eCRF may lower mortality risk to a greater extent in females through anti-inflammatory pathways.^49^ Finally, from a behavioral perspective, women generally engage in lower levels of habitual physical activity than men, so changes in eCRF may more sensitively reflect lifestyle modifications in women and thus mediate a larger proportion of the mortality risk. Sex-specific mediation analysis reveals that improving exercise capacity (e.g., aerobic training) is the priority for counteracting CTI-FI risk in women, whereas directly reducing CTI-FI is more critical in men.

## Limitations

First, the FI construction differed between the two cohorts: NHANES used 49 deficits whereas CHARLS used 32 deficits, and some item definitions were not completely identical. Although we calculated FI separately for each cohort, cross-cohort comparisons of CTI-FI should be interpreted with caution. Second, the eCRF estimation formula was derived from a Western population and its application to the Chinese population may introduce calibration bias; future validation using Chinese population-based eCRF equations is needed. Third, the combined model outperformed the FRS for CVM, but this advantage was only assessed in the NHANES cohort using 10-fold cross-validation and could not be externally validated (CHARLS had only ACM data). Fourth, although we used E-values and multiple sensitivity analyses (including exclusion of short follow-up, landmark analyses, LASSO-based variable selection, and Fine-Gray competing risk models) to address measured and unmeasured confounding, residual confounding inherent to observational study designs cannot be completely ruled out. Future prospective cohort studies and randomized trials are warranted to further validate the clinical value of our findings.

## Conclusions

In summary, CTI-FI and eCRF are independently and jointly associated with mortality, with bidirectional mediation and notable sex heterogeneity. Adding both biomarkers to a basic risk model significantly improved predictive discrimination. External validation in the CHARLS cohort confirmed generalizability for ACM. These findings position CTI-FI and eCRF as complementary, modifiable predictors for early risk stratification and management of CKM syndrome.

## Abbreviations

CKM: cardiovascular-kidney-metabolic syndrome
CTI-FI: C-reactive protein-triglyceride glucose index-frailty index
eCRF: non-exercise estimated cardiorespiratory fitness
TyG-FI: triglyceride-glucose index combined with frailty index
CHG-FI: combined cholesterol, high-density lipoprotein, glucose, and frailty index
Fine-Gray: Fine-Gray subdistribution hazard

## Data Availability

The raw data for the tables and figures in this article were obtained from the public databases NHANES and CHARLS.

## Acknowledgements

We are grateful to the staff and participants of the NHANES and CHARLS databases for their contributions. Conceptualization: Jie An, Dandan Yang, Jie Li and Qiang She; Investigation: Jie An, Qianhua Feng and Ya Luo; Formal analysis: Jie An, Qianhua Feng and Dandan Yang; Writing—original draft and review & editing: Jie An, Qianhua Feng, Jie Li and Qiang She; Supervision: Ya Luo, Meng Yu and Min Xu; All authors have read and agreed to the published version of the manuscript.

## Clinical Perspective

### What is new?

Per 1-unit increase was associated with a 44% higher risk of ACM and a 54% higher risk of CVM. Per 1-MET higher eCRF, ACM and CVM risks were reduced by 10% and 18%, respectively.

Compared with the Low CTI-FI + High eCRF group, the High CTI-FI + Low eCRF group had a 2.74-fold higher risk of ACM and a 5.04-fold higher risk of CVM. Bidirectional mediation: eCRF explained 16-24% of CTI-FI-related mortality risk, while CTI-FI explained 24-32% of eCRF ’ s survival benefit. Dose-response and mediation patterns differed by sex and population.

### What Are the Clinical Implications?

This study introduces two non-exercise, easily ascertainable indices — CTI-FI and eCRF—for early risk stratification in adults with CKM stages 0-3. The bidirectional mediation highlights that improving either metabolic-inflammatory status (CTI-FI) or cardiorespiratory fitness (eCRF) could partially counteract the adverse effect of the other, informing targeted lifestyle and pharmacological interventions.

